# Pharmacogenomic architecture of antihypertensive switching implicates neurotensin-NTSR1 signaling in ACE inhibitor-induced cough

**DOI:** 10.64898/2026.01.26.26344813

**Authors:** Felix Vaura, Kristi Krebs, Tuomo Kiiskinen, Joel Rämö, Max Tamlander, FinnGen, Estonian Biobank research team, Simone Rubinacci, Lili Milani, Samuli Ripatti

## Abstract

Early failure of antihypertensive medication treatment affects one in four patients, but the underlying mechanisms are poorly understood. We aimed to identify genetic determinants of antihypertensive treatment failure within the first year.

Using longitudinal medication data from >400,000 genotyped antihypertensive medication users across 3 cohorts (FinnGen, the UK Biobank, and the Estonian Biobank), we classified short-term antihypertensive use trajectories as Continue, Switch, or Discontinue. We performed genome-wide association studies of switching across five major medication classes, followed by replication and downstream analyses linking genetic loci to medication-specific adverse events and clinical outcomes.

We identified 14 genome-wide significant loci for switching from angiotensin-converting enzyme inhibitors (ACEI) and dihydropyridine calcium channel blockers (dCCB) to other antihypertensive medications. For ACEI switching, multiple independent signals converged on the neurotensin-NTSR1 pathway, including a 320-fold Finnish-enriched protective missense variant in the neurotensin receptor gene *NTSR1* (rs148569146 [G301R], allele frequency [AF] 1.4%, odds ratio [OR] 0.49, P=3.3×10^−43^) and a variant near *RASSF9* (rs181941187, AF=9.0%, OR=0.74, P=1.2×10^−49^) tagging the neurotensin gene *NTS*. In drug-gene interaction analyses, *NTSR1* G301R was associated with reduced ACEI-induced cough risk (OR=0.39, P=8.1×10^−4^). The dCCB switching locus at *CYP3A43* was in near-complete linkage (r^2^=0.99) with the functional *CYP3A4*22* allele (rs35599367, AF=3.4%, OR=1.23, P=6.1×10^−10^).

Our findings extend the bradykinin hypothesis of ACEI-induced cough by implicating neurotensin-NTSR1 signaling. We additionally characterize *CYP3A4*22* as a mechanistically supported predictor of dCCB intolerance. Longitudinal medication trajectories offer a rich framework for pharmacogenetic discovery with potential for future genotype-informed antihypertensive medication selection.

**Graphical abstract:** 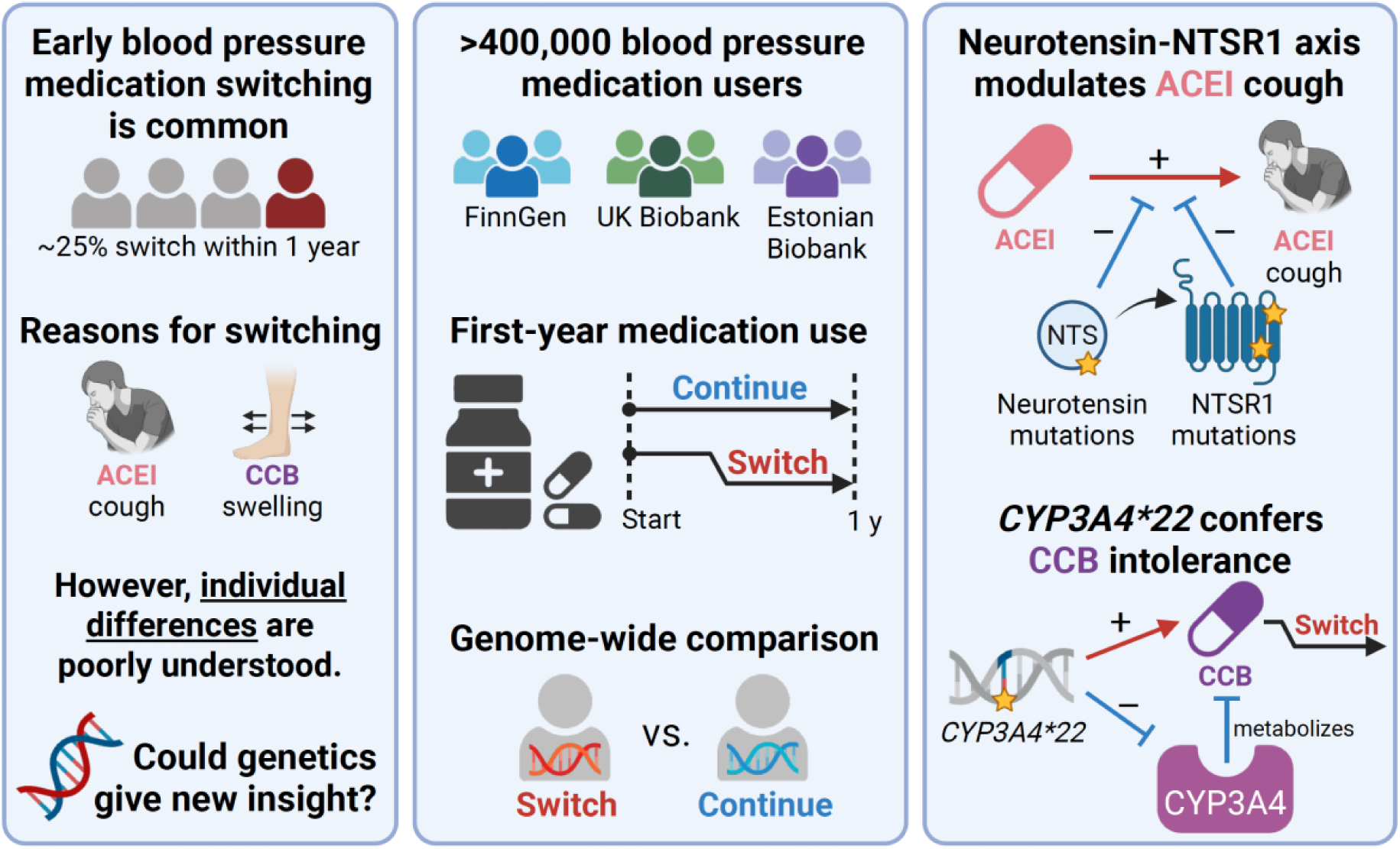

## Introduction

High blood pressure (hypertension) is the leading metabolic risk factor for global disease burden,[1] affecting over a billion people worldwide despite the availability of multiple effective medication classes.[2] Choosing the right antihypertensive medication is challenging, with one in four patients switching treatment within the first year.[3] The biological basis of this variation in treatment trajectories remains poorly understood, and few tools exist to guide personalized antihypertensive medication selection.

Genetic variability is showing promise for personalizing hypertension care,[4] and pharmacogenetic studies have indicated that common genetic variants influence both antihypertensive medication efficacy and tolerability.[5] However, most studies have been small and underpowered for genome-wide discovery of underlying genetic loci.[5] Moreover, conventional laboratory-derived medication response phenotypes, such as changes in plasma concentrations, may not translate to real-world treatment trajectories such as medication switching or discontinuation.

The integration of genomic data and longitudinal health records has opened new avenues for large-scale pharmacogenetic discovery. Recent meta-analyses have demonstrated that angiotensin-converting enzyme inhibitor (ACEI) switching has a polygenic basis in non-coding variation,[6,7] while well-known blood pressure loci have been shown to cumulatively influence both long-term antihypertensive use and early discontinuation.[8,9] Despite these advances, little progress has been made toward understanding the biological processes driving antihypertensive treatment failure.

We aimed to identify novel genetic variants underlying short-term antihypertensive treatment failure. To do this, we integrated genome-wide profiles with longitudinal medication data to construct first-year antihypertensive use trajectories in more than 400,000 individuals from FinnGen, the UK Biobank (UKB), and the Estonian Biobank (EstBB).

## Methods

### Study samples

We performed a two-stage study combining genetic discovery in FinnGen and replication in UKB and EstBB.[10–12] FinnGen data freeze 12 comprises 520,210 participants genotyped and imputed to 21.3 million genetic variants and linked to national electronic health registers with simultaneous medical diagnosis code and medication purchase follow-up spanning 1995–2022. UKB includes 487,180 genotyped participants, of whom 221,863 have primary care medication prescription follow-up spanning 1990–2017. EstBB data freeze 2024v1 comprises 211,259 genotyped participants linked to national health records, including dispensed medications, spanning 2004–2024.

### Medication data

In FinnGen and EstBB, we inferred medication use from outpatient medication purchases, while in UKB we used primary care prescriptions. Medication purchase data were obtained from the Social Insurance Institution of Finland in FinnGen, the Estonian Health Insurance Fund in EstBB, and the National Health Service in UKB. We considered five major antihypertensive medication classes defined by anatomical therapeutic chemical (ATC) codes: thiazide diuretics (THZ, ATC C03A*), selective β-blockers (sBB, ATC C07AB*), dihydropyridine calcium channel blockers (dCCB, ATC C08CA*), angiotensin-converting enzyme inhibitors (ACEI, ATC C09A*), and angiotensin 2 receptor blockers (ARB, ATC C09C*).[2] Because UKB prescriptions were coded with British National Formulary, dictionary of medicines and devices, and read version 2 codes, we mapped them to ATC codes using a publicly available pipeline by Sadler et al.[13]

### Classification of antihypertensive use patterns

We classified short-term medication patterns within each of the five major antihypertensive classes into three categories: Continue, Switch, and Discontinue (**Supplementary Figure 1**). For a given antihypertensive class (the index class), we first identified the participant’s first-ever purchase or prescription of any medication in that class at time T_F_ (the index medication). We then identified the last index medication purchase or prescription occurring within one year of T_F_ at time T_L_.

We defined two index class-specific windows: the co-prescription window (T_F_ – 1 year, T_L_] and the classification window (T_L_, T_L_ + 1 year] (**Supplementary Figure 1**). Participants were classified as Continue if the classification window contained the index medication. Those not meeting this criterion were classified as Switch if the classification window contained an antihypertensive medication absent from the co-prescription window. All remaining participants were classified as Discontinue (**Supplementary Figure 1**).

Participants with <1 year of electronic health register follow-up before T_F_, <2 years of follow-up after T_F_, or age <18 years at T_F_ were excluded from the corresponding index class analysis.

### Disease endpoints

For the medical code enrichment analysis, we used the International Classification of Diseases, 10th revision (ICD-10) and the International Classification of Primary Care, 2nd edition (ICPC-2) codes. For phenome-wide association studies (PheWAS), we used 2,469 curated clinical endpoints from FinnGen data freeze 12, which integrate several national and international coding systems. FinnGen clinical endpoint definitions are publicly accessible at https://www.finngen.fi/en/researchers/clinical-endpoints.

### Genotyping and imputation

We used imputed genotype data from FinnGen, UKB, and EstBB, as well as whole-exome sequencing data from UKB. FinnGen samples were genotyped with Illumina and Affymetrix arrays and imputed against the SISu v4.2 reference panel based on 8,554 whole genome-sequenced Finns. Genotype data quality control (QC) involved sample-wise (call rate, heterozygosity, sex discordance) and variant-wise (call rate, Hardy-Weinberg equilibrium P value, minor allele count) steps described in detail by Kurki et al.[10] UKB genotype data was genotyped with custom Affymetrix arrays and imputed against the Trans-Omics for Precision Medicine (TOPMed) Program imputation panel R2 (UKB field 21007).[14] UKB genotype data QC involved several steps with careful handling of diverse ancestry and additional QC in the TOPMed imputation server prior to imputation.[11,15] UKB whole-exome sequencing data (UKB field 23159) was generated at the Regeneron Genetics Center with sequencing, variant calling, and quality control previously described in detail.[16] EstBB genotype data was genotyped with Illumina Arrays and imputed against a population-specific reference panel consisting of 2,695 whole genome-sequenced Estonians.[17] Genotype data quality control involved sample-wise (call rate, heterozygosity, sex discordance) and variant-wise (call rate, Hardy-Weinberg equilibrium P value, minor allele frequency) steps described in detail by Abner et al.[17] Our analysis excluded variants with INFO score <0.6 and included individuals from major ancestry groups in each biobank: Finnish in FinnGen, white British in UKB, and European in EstBB.

### Discovery GWAS

For each of the five major antihypertensive classes in FinnGen, we performed a GWAS comparing the Switch group (cases) with the Continue group (controls). To assess the genetic overlap between switching and discontinuing phenomena, we additionally performed GWASs comparing the Discontinue group (cases) with the Continue group (controls). We performed association testing with regenie version 2.2.4, adjusting for age, age squared, genetic sex, first 10 genetic principal components, and genotyping batch (dummy-coded). We considered P<5×10^−8^ as genome-wide significant.

### Fine-mapping and annotation

To identify likely causal variants, we fine-mapped 3 Mb regions around genome-wide significant loci of Switch phenotypes using SuSiE version 0.9.2 assuming up to 10 causal variants per region.[18] In-sample dosages were computed with LDstore2.[19] For downstream analyses, we retained lead variants from high-quality credible sets (CS), defined as log10 Bayes factor >2 for the CS and minimum linkage disequilibrium (LD) r^2^ >0.20 between CS variants. We annotated variant consequences using the Variant Effect Predictor version 104 and estimated Finnish enrichment compared to non-Finnish Europeans using gnomAD version 4.0.[20,21]

### Replication and coding-variant meta-analysis

We first replicated FinnGen CS lead variants in a UKB-EstBB meta-analysis using imputed genotype data. Full replication required concordant effect direction and P<0.05/k where k is the number of high-quality CS lead variants (Bonferroni correction). We performed association testing in UKB and EstBB with regenie version 2.1.0, adjusting for age, age squared, sex, and the first 10 genetic principal components.

Next, to prioritize mechanistically plausible genetic variants and to boost statistical power, we performed a protein-coding variant meta-analysis across FinnGen, UKB (exome data), and EstBB. We defined protein-coding variants as those in the following categories: stop gained, frameshift, splice acceptor, splice donor, in-frame insertion, in-frame deletion, missense, start lost, or stop lost. To decrease type I error, we imposed a stricter genome-wide significance threshold of P<5×10^−9^ in addition to concordant effect direction in at least two out of three cohorts. We performed fixed-effect inverse variance-weighted meta-analyses with METAL version 2020-05-05.[22]

### Medical code enrichment analysis and variant annotation

To investigate whether our Switch phenotypes capture known adverse events underlying antihypertensive treatment failure,[2] we performed an enrichment analysis of medical codes (ICD-10 and ICPC-2) in FinnGen. For each major antihypertensive class, we predicted Switch group status with medical code incidence using logistic regression, as has been done before.[23] Medical code incidence was ascertained between the first purchase date and either the switch date (Switch group) or the end of the first year (Continue group). For each medical code, we excluded individuals with prevalent codes at baseline (first purchase date) and required each medical code to have at least 5 cases in both the Switch group and the Continue group. We adjusted the logistic regression model by age, sex, and length of follow-up, and considered Bonferroni-corrected P as statistically significant. To interrogate CS lead variants on their associations to main antihypertensive class-specific adverse events, we additionally conducted adverse event GWASs within all participants combined, including the Discontinue group. We defined cases as those who had at least one recorded adverse event-related ICD-10 or ICPC-2 code within 1 year of baseline.

### Phenome-wide association study

To characterize CS lead variants from a broad clinical perspective, we used phenome-wide association study (PheWAS) results derived from 2,469 curated FinnGen endpoints as well as 3 blood pressure measurements from the largest blood pressure GWAS to date.[24] We considered a Bonferroni-corrected P<0.05/2,475 as statistically significant.

### Heritability and genetic overlap

We estimated the SNP-based heritability of antihypertensive switching and discontinuation using linkage disequilibrium score regression (LDSC, version 1.0.1) applied to FinnGen discovery GWAS results.[25] To assess genetic overlap within and between Switch and Discontinue traits, we used two complementary approaches. First, we computed all pairwise genetic correlations with LDSC. Second, because discontinuation can be confounded by behavioural traits unrelated to medication response, we hypothesized that CS lead variant effects from the Switch GWAS would be proportionally attenuated in the corresponding Discontinue GWAS. To test this, we fitted a single-slope model

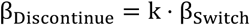

using linemodels version 0.5.0 and estimated posterior probabilities for three models: slope = 0, k, and 1.[26]

### Statistical analyses

To estimate drug-gene interactions between antihypertensive medication exposure and Switch GWAS hits on adverse events, we used a self-controlled risk interval design with fixed-effects conditional logistic regression.[27] For each participant, we defined two 1-year risk intervals relative to the first medication purchase at *t*_0_: a pre-exposure interval [*t*_0_ − 1.5 years, *t*_0_ − 0.5 years] and a post-exposure interval [*t*_0_, *t*_0_ + 1 year], separated by a 6-month washout period. Event incidence was coded as 0 or 1 in each interval using ICD-10 and ICPC-2 diagnoses. We fit the model

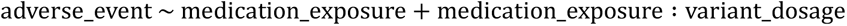

to estimate the drug-gene interaction term (R package *survival*, function clogit). The self-controlled design inherently adjusts for main effects of all time-invariant confounders such as co-morbidities, co-prescriptions, environmental variables, and relatedness.[27]

### Sensitivity analyses

To investigate the effect of dose-adjustment and adherence on the discovery GWAS results in FinnGen, we performed two sensitivity analyses for all Switch GWASs with genome-wide significant hits: (1) additionally adjusting by the first purchase dose measured in the World Health Organization’s Defined Daily Dose units (rank normalised), and (2) requiring consecutive index medication and switch medication purchases (**Supplementary Figure 1**) to be at most 180 days apart. To compare effect size estimates of CS lead variants before and after these adjustments, we plotted them for visual inspection.

### Ethical approval

Study subjects in FinnGen provided informed consent for biobank research, based on the Finnish Biobank Act. Alternatively, separate research cohorts, collected prior the Finnish Biobank Act came into effect (in September 2013) and start of FinnGen (August 2017), were collected based on study-specific consents and later transferred to the Finnish biobanks after approval by Fimea (Finnish Medicines Agency), the National Supervisory Authority for Welfare and Health. Recruitment protocols followed the biobank protocols approved by Fimea. The Coordinating Ethics Committee of the Hospital District of Helsinki and Uusimaa (HUS) statement number for the FinnGen study is Nr HUS/990/2017.

The FinnGen study is approved by Finnish Institute for Health and Welfare (permit numbers: THL/2031/6.02.00/2017, THL/1101/5.05.00/2017, THL/341/6.02.00/2018, THL/2222/6.02.00/2018, THL/283/6.02.00/2019, THL/1721/5.05.00/2019 and THL/1524/5.05.00/2020), Digital and population data service agency (permit numbers: VRK43431/2017-3, VRK/6909/2018-3, VRK/4415/2019-3), the Social Insurance Institution (permit numbers: KELA 58/522/2017, KELA 131/522/2018, KELA 70/522/2019, KELA 98/522/2019, KELA 134/522/2019, KELA 138/522/2019, KELA 2/522/2020, KELA 16/522/2020), Findata permit numbers THL/2364/14.02/2020, THL/4055/14.06.00/2020, THL/3433/14.06.00/2020, THL/4432/14.06/2020, THL/5189/14.06/2020, THL/5894/14.06.00/2020, THL/6619/14.06.00/2020, THL/209/14.06.00/2021, THL/688/14.06.00/2021, THL/1284/14.06.00/2021, THL/1965/14.06.00/2021, THL/5546/14.02.00/2020, THL/2658/14.06.00/2021, THL/4235/14.06.00/2021, Statistics Finland (permit numbers: TK-53-1041-17 and TK/143/07.03.00/2020 (earlier TK-53-90-20) TK/1735/07.03.00/2021, TK/3112/07.03.00/2021) and Finnish Registry for Kidney Diseases permission/extract from the meeting minutes on 4th July 2019. The Biobank Access Decisions for FinnGen samples and data utilized in FinnGen Data Freeze 12 include: THL Biobank BB2017_55, BB2017_111, BB2018_19, BB_2018_34, BB_2018_67, BB2018_71, BB2019_7, BB2019_8, BB2019_26, BB2020_1, BB2021_65, Finnish Red Cross Blood Service Biobank 7.12.2017, Helsinki Biobank HUS/359/2017, HUS/248/2020, HUS/430/2021 §28, §29, HUS/150/2022 §12, §13, §14, §15, §16, §17, §18, §23, §58, §59, HUS/128/2023 §18, Auria Biobank AB17-5154 and amendment #1 (August 17 2020) and amendments BB_2021-0140, BB_2021-0156 (August 26 2021, Feb 2 2022), BB_2021-0169, BB_2021-0179, BB_2021-0161, AB20-5926 and amendment #1 (April 23 2020) and it’s modifications (Sep 22 2021), BB_2022-0262, BB_2022-0256, Biobank Borealis of Northern Finland_2017_1013, 2021_5010, 2021_5010 Amendment, 2021_5018, 2021_5018 Amendment, 2021_5015, 2021_5015 Amendment, 2021_5015 Amendment_2, 2021_5023, 2021_5023 Amendment, 2021_5023 Amendment_2, 2021_5017, 2021_5017 Amendment, 2022_6001, 2022_6001 Amendment, 2022_6006 Amendment, 2022_6006 Amendment, 2022_6006 Amendment_2, BB22-0067, 2022_0262, 2022_0262 Amendment, Biobank of Eastern Finland 1186/2018 and amendment 22§/2020, 53§/2021, 13§/2022, 14§/2022, 15§/2022, 27§/2022, 28§/2022, 29§/2022, 33§/2022, 35§/2022, 36§/2022, 37§/2022, 39§/2022, 7§/2023, 32§/2023, 33§/2023, 34§/2023, 35§/2023, 36§/2023, 37§/2023, 38§/2023, 39§/2023, 40§/2023, 41§/2023, Finnish Clinical Biobank Tampere MH0004 and amendments (21.02.2020 & 06.10.2020), BB2021-0140 8§/2021, 9§/2021, §9/2022, §10/2022, §12/2022, 13§/2022, §20/2022, §21/2022, §22/2022, §23/2022, 28§/2022, 29§/2022, 30§/2022, 31§/2022, 32§/2022, 38§/2022, 40§/2022, 42§/2022, 1§/2023, Central Finland Biobank 1-2017, BB_2021-0161, BB_2021-0169, BB_2021-0179, BB_2021-0170, BB_2022-0256, BB_2022-0262, BB22-0067, Decision allowing to continue data processing until 31st Aug 2024 for projects: BB_2021-0179, BB22-0067,BB_2022-0262, BB_2021-0170, BB_2021-0164, BB_2021-0161, and BB_2021-0169, and Terveystalo Biobank STB 2018001 and amendment 25th Aug 2020, Finnish Hematological Registry and Clinical Biobank decision 18th June 2021, Arctic biobank P0844: ARC_2021_1001.

The activities of the EstBB are regulated by the Human Genes Research Act, which was adopted in 2000 specifically for the operations of the EstBB. Individual level data analysis in the EstBB was carried out under ethical approval “1.1-12/624” from the Estonian Committee on Bioethics and Human Research (Estonian Ministry of Social Affairs), using data according to release application “6-7/GI/29470” from the Estonian Biobank.

## Results

### Sample characteristics and switch trajectories

In FinnGen, we identified 256,561 participants (mean age 56.2, 51.7% female) who initiated treatment with one or more major antihypertensive classes: ACEI, THZ, dCCB, sBB, or ARB. All participants had >1 year of electronic health register follow-up prior to initiation and >2 years thereafter. Cohort-specific characteristics are summarized in **Supplementary Table 1**.

Rate of antihypertensive switching ranged from 11% in ARB to 32% in ACEI, and the median switch time was 4–5 months across antihypertensive classes (**Supplementary Figure 2a**). In contrast, more than 50% of discontinuation happened before the second purchase (**Supplementary Figure 2b**). Switching patterns varied across antihypertensive classes, with the ACEI-to-ARB switch having the highest rate of 57% of ACEI switches (**Supplementary Figure 3**).

### Genetic discovery and replication of antihypertensive switching

We first tested for genetic associations genome-wide for switching away from antihypertensive medication classes. We found 14 genome-wide significant loci for switching away from ACEI and dCCB to any other antihypertensive medication. We observed no genome-wide loci for sBB, THZ, or ARB switching.

The ACEI switching GWAS yielded 13 genome-wide loci that fine-mapped to 21 high-quality credible sets, including four novel loci at *OSBPL10*, *SYBU*, *CACNA1H*, and *SHISAL1* (**Figure 1**, **Table 1**). Notably, we identified two missense variants in our credible sets for ACEI switching: a protective 320-fold Finnish-enriched variant p.Gly301Arg in *NTSR1* (rs148569146-C, multiallelic site, allele frequency [AF] =1.4%, odds ratio [OR] =0.49, P=3.3×10^−43^) and risk-increasing p.Val664Ala in *CACNA1H* (rs4984636-C, AF=32%, OR=1.07, P=9.8×10^−9^). The *NTSR1* missense variant formed a single-variant CS and localizes to the intracellular allosteric site of neurotensin receptor 1 (NTSR1, **Figure 2**, **Supplementary Table 2**). The *NTSR1* locus contained another single-variant CS with rs6062847-T, a previously identified downstream gene variant for *LINC00686*, which is independent from the *NTSR1* missense variant (LD r^2^=0.002; **Figure 3**). Moreover, the overall strongest GWAS signal was a CS lead variant in chromosome 12 near *RASSF9* (rs181941187-G, AF=9.0%, 40-fold Finnish-enriched, OR=0.74, P=1.2×10^−49^), which was in moderate LD (r^2^ 0.4–0.6) with a cluster of variants at *NTS* – the gene encoding neurotensin i.e. the endogenous ligand of NTSR1 (**Figure 3**).

**Figure 1.**
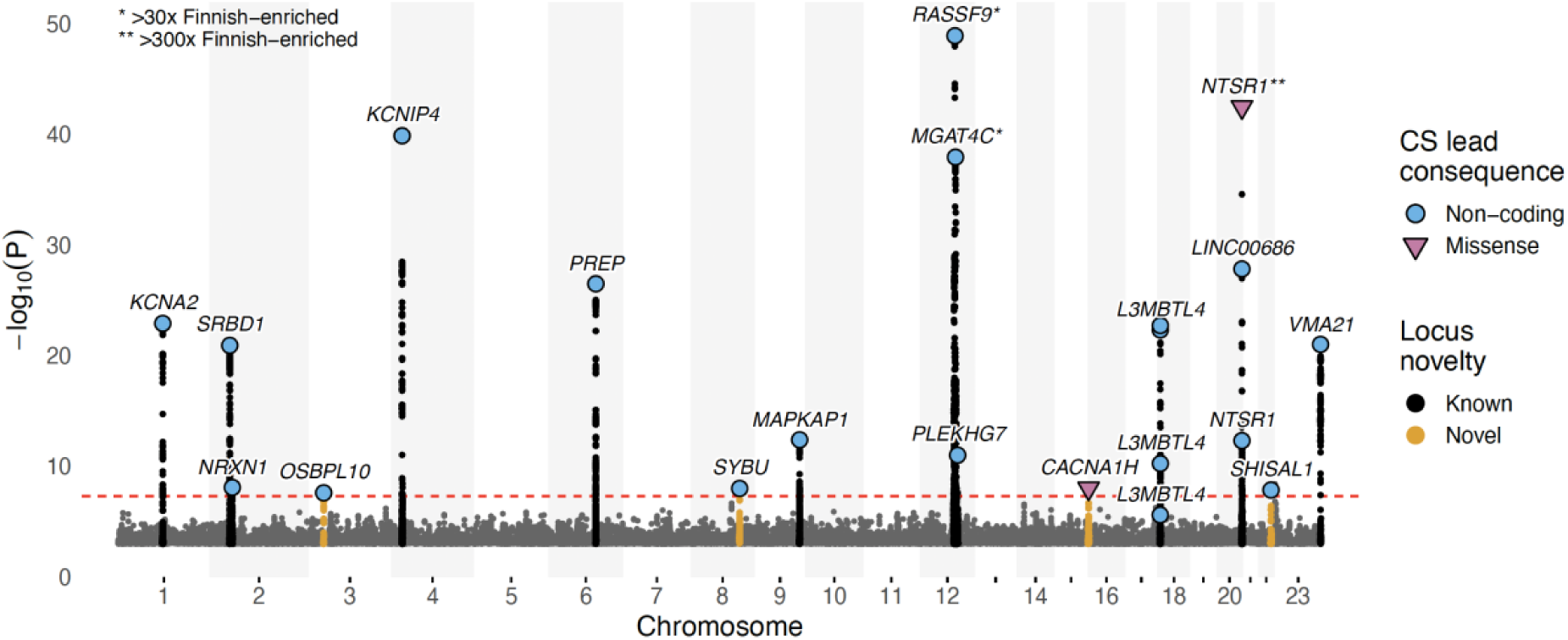
Genome-wide association results for ACEI switching in FinnGen. Manhattan plot showing genome-wide significant credible set (CS) lead variants for switching from ACEI to any other antihypertensive class in FinnGen (23,858 cases, 56,273 controls). Missense (purple) and non-coding (blue) CS lead variants are indicated by colour. Novel loci are highlighted in yellow. Finnish-enriched variants are marked by asterisks. ACEI, angiotensin-converting enzyme inhibitor; CS, credible set.

**Figure 2.**
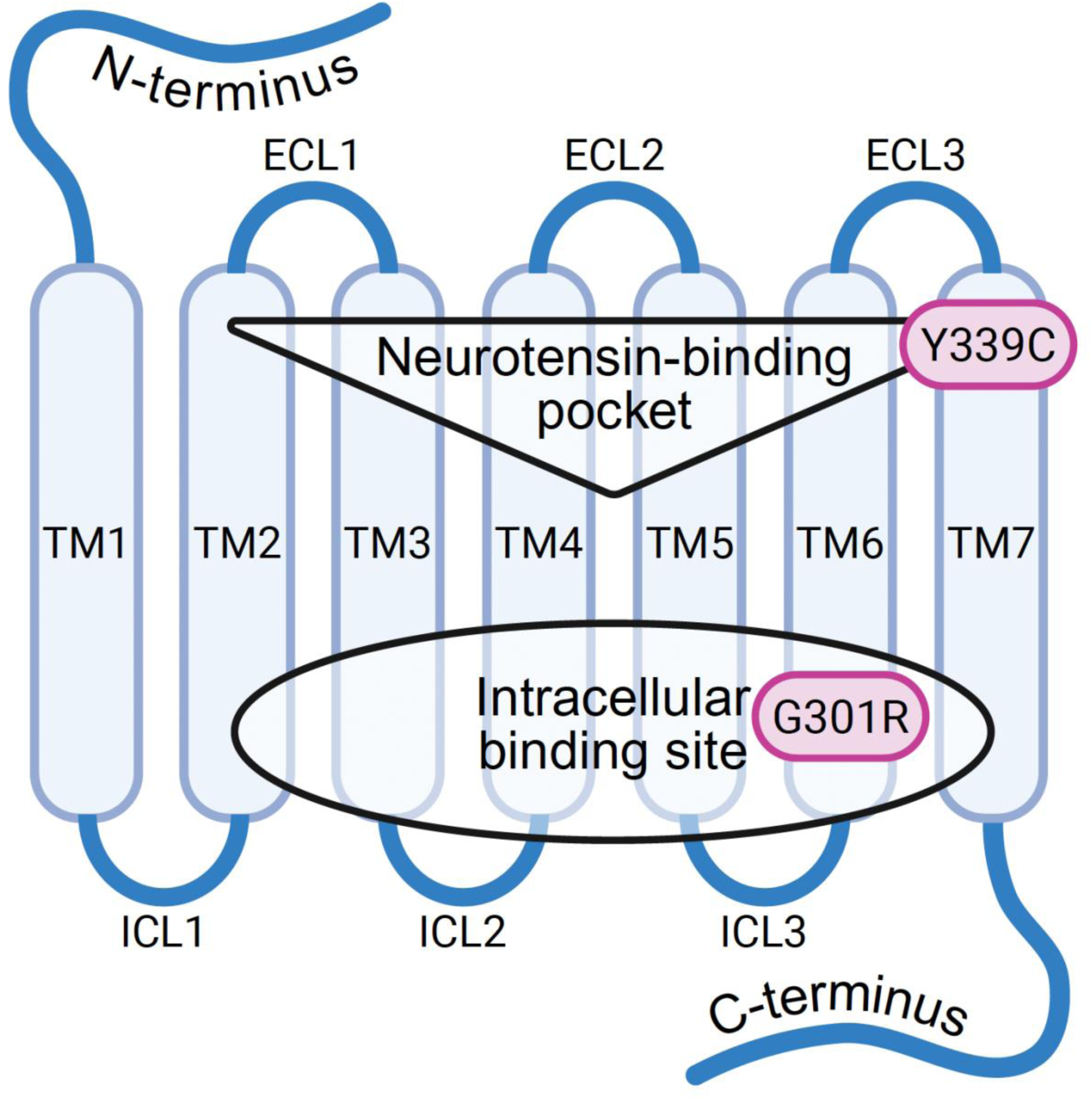
Schematic structure of the G protein-coupled receptor NTSR1 showing the locations of two ACEI Switch missense variants. The 320-fold Finnish-enriched p.Gly301Arg (G301R) localizes to the intracellular binding site on TM6, while the 0.08-fold Finnish-depleted p.Tyr339Cys (Y339C) localizes to the neurotensin-binding pocket on TM7. ACEI, angiotensin-converting enzyme inhibitor; ECL, extracellular loop; ICL, intracellular loop; NTSR1, neurotensin receptor 1; TM, transmembrane helix. Created with BioRender.com.

**Figure 3.**
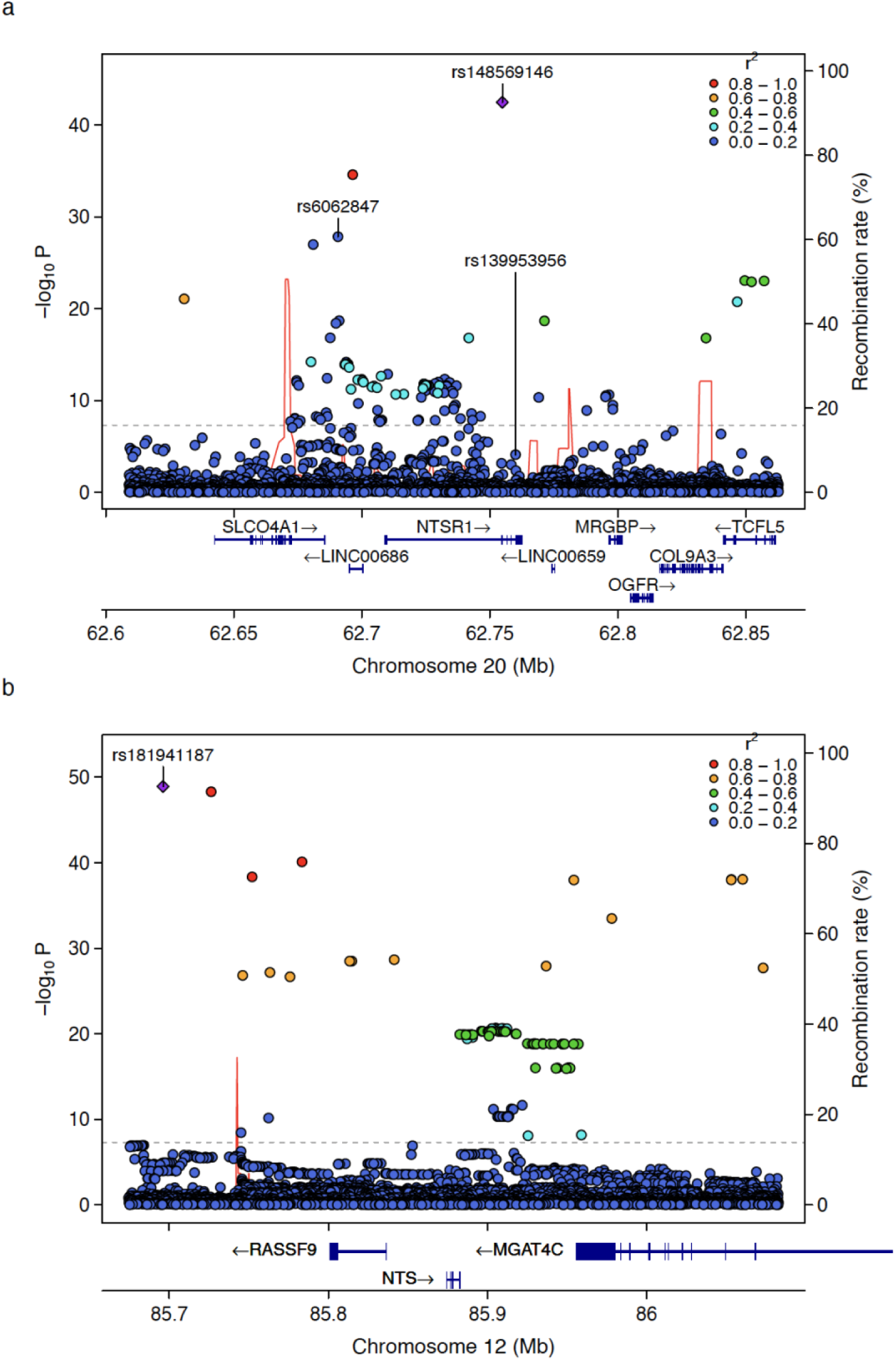
Converging genetic evidence implicates the neurotensin-NTSR1 axis in ACEI intolerance. (a) Fine-mapping the *NTSR1* locus on chromosome 20 reveals two independent credible set lead variants rs148569146-C (*NTSR1*, p.Gly301Arg) and rs6062847-T (*LINC00686*), as well as an independent coding variant meta-analysis signal rs139953956-G (*NTSR1*, p.Tyr339Cys). (b) Fine-mapping of the *RASSF9*-*NTS* locus on chromosome 12 shows the top GWAS hit rs181941187-G tagging a cluster of variants near the neurotensin gene *NTS*. We display linkage disequilibrium r^2^ (colour scale) relative to the lead variants (purple diamond), and the recombination rate (red line). ACEI, angiotensin-converting enzyme inhibitor; NTSR1, neurotensin receptor 1.

**Table 1.**
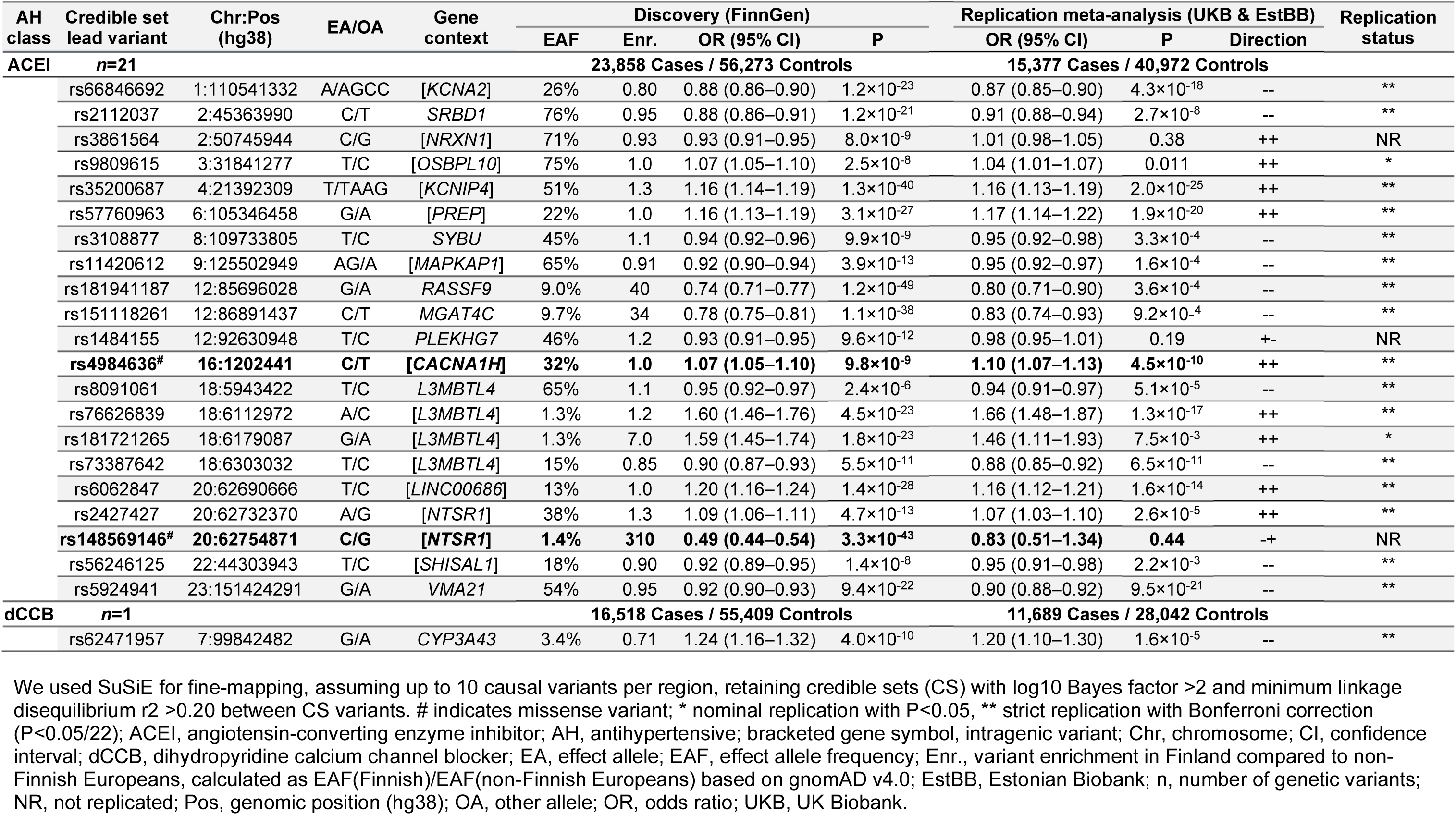
Discovery and replication summary statistics for fine-mapped lead variants.

We used SuSiE for fine-mapping, assuming up to 10 causal variants per region, retaining credible sets (CS) with log10 Bayes factor >2 and minimum linkage disequilibrium r2 >0.20 between CS variants. # indicates missense variant; * nominal replication with P<0.05, ** strict replication with Bonferroni correction (P<0.05/22); ACEI, angiotensin-converting enzyme inhibitor; AH, antihypertensive; bracketed gene symbol, intragenic variant; Chr, chromosome; CI, confidence interval; dCCB, dihydropyridine calcium channel blocker; EA, effect allele; EAF, effect allele frequency; Enr., variant enrichment in Finland compared to non-Finnish Europeans, calculated as EAF(Finnish)/EAF(non-Finnish Europeans) based on gnomAD v4.0; EstBB, Estonian Biobank; n, number of genetic variants; NR, not replicated; Pos, genomic position (hg38); OA, other allele; OR, odds ratio; UKB, UK Biobank.

The dCCB switching GWAS yielded a single genome-wide significant locus at *CYP3A43* whose lead variant rs62471957-G is in near-complete LD (r^2^=0.99) with the recently characterised CYP3A4 enzyme star allele *CYP3A4*22* (rs35599367-A, AF=3.4%, OR=1.23, P=6.1×10^−10^) (**Supplementary Table 2**).

We additionally observed two non-coding genome-wide significant signals for ACEI discontinuation: rs77832029 near *KCNIP4* (AF=7.4%, OR=1.16, P=4.2×10^−9^) and rs2322536 near *L3MBTL4* (AF=1.6%, OR=1.34, P=2.8×10^−8^).

### Heritabilities, genetic correlations, and effect size attenuation

We estimated the SNP heritability for ACEI switching to be 6.8% (95% CI, 3.4–10.3) with other Switch and Discontinue phenotypes displaying lower heritability estimates in the range 0–3% (**Figure 4a, Supplementary Tables 3 and 4**). After correcting for multiple testing across 45 phenotype pairs, we estimated several Discontinue-Discontinue phenotype pairs to be highly correlated (**Figure 4b**).

**Figure 4.**
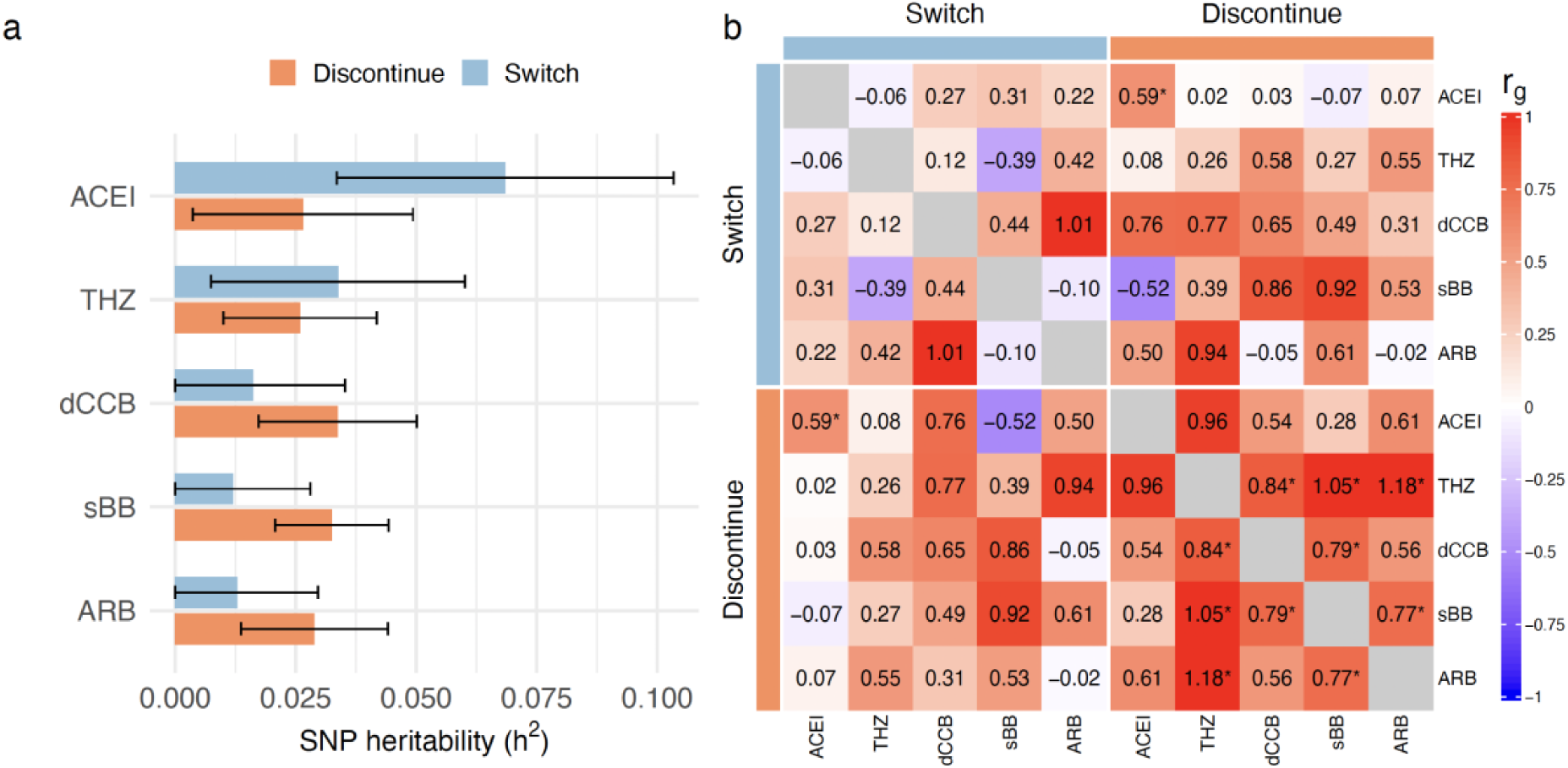
SNP-based heritability (h^2^) and genetic correlations (r_g_) of antihypertensive use phenotypes across five major antihypertensive classes. (a) h^2^ estimates on the liability scale for Switch (blue) and Discontinue (orange) phenotypes with black error bars representing 95% confidence intervals. (b) r_g_ heatmap for Switch and Discontinue phenotypes, with colour denoting positive (red) and negative (blue) correlations and asterisks (*) denoting statistically significance (P<0.05/45) after Bonferroni correction over 45 phenotype pairs. We used linkage disequilibrium score regression to estimate h^2^ and r_g_. ACEI, angiotensin-converting enzyme inhibitor; ARB, angiotensin receptor 2 blocker; dCCB, dihydropyridine calcium channel blocker; sBB, selective beta blocker; SNP, single nucleotide polymorphism; THZ, thiazide diuretic.

Switch-Discontinue correlation point estimates were overall positive (except for ACEI-sBB), but only the ACEI-ACEI pair remained statistically significant with r_g_=0.59 (95% CI, 0.26–0.92). Switch-Switch correlation point estimates were moderate to low (**Figure 4b**). In FinnGen, ACEI Switch variant effect sizes were consistently attenuated in the Discontinue phenotype by ∼0.4, while ACEI Discontinue variants showed no attenuation in the Switch phenotype (**Supplementary Figure 4**).

### Replication and coding variant meta-analysis

After meta-analysing UKB and EstBB (15,377 cases and 40,972 controls, **Supplementary Table 1**) using inverse variance-weighted meta-analysis, we replicated all 13 ACEI switching loci nominally (P <0.05), with full (P<0.05/22) replication for 3/4 novel and 8/9 previously known loci (**Table 1**, **Supplementary Table 5**). We also replicated the dCCB locus at *CYP3A43*, including the star allele *CYP3A4*22* (OR=1.20, P=1.3×10^−5^). In the coding variant meta-analysis combining FinnGen, UKB, and EstBB (39,235 cases and 97,245 controls) we identified additional coding variants for ACEI switching in *PREP*, *CACNA1H*, and *NTSR1*, as well as one coding variant for dCCB switching in *CYP3A4* (**Table 2**). In sensitivity analyses, effect size estimates for all ACEI and dCCB Switch CS lead variants were robust to dose-adjustment and stricter adherence definitions (**Supplementary Figures 5 and 6**).

**Table 2.**
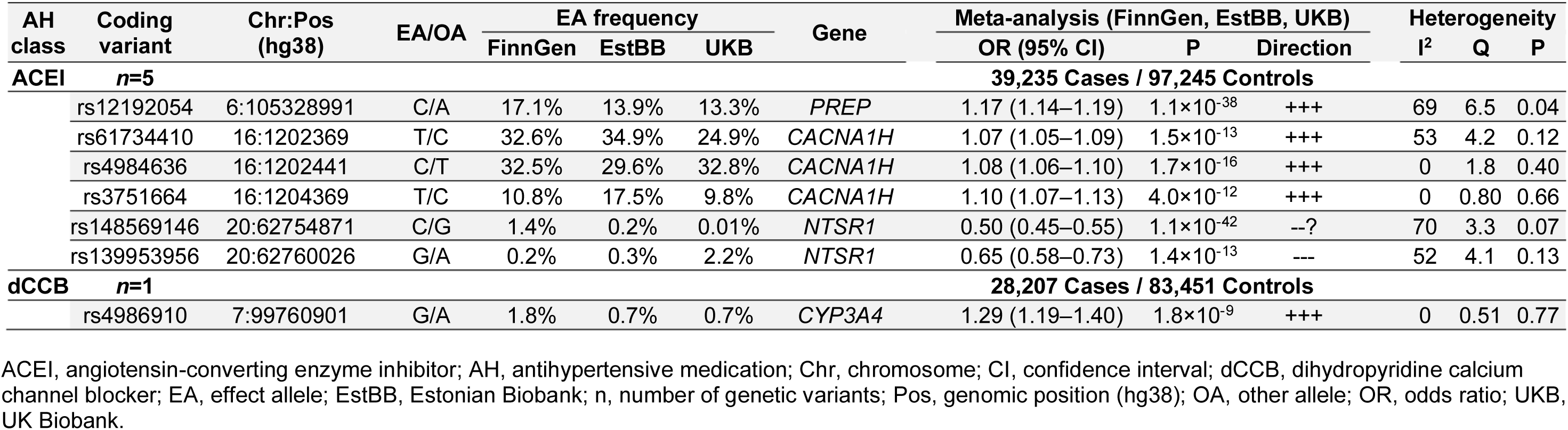
Coding variant meta-analysis results across FinnGen, the UK Biobank, and the Estonian Biobank.

### Medical code enrichment analysis

We observed enrichment of several known adverse events in the Switch group compared with the Continue group: cough and angioedema in ACEI switching, hyponatremia and hypokalemia in THZ switching, localized oedema in dCCB switching, and bradycardia in sBB switching (P<1.3×10^−5^; **Supplementary Table 6**). The Switch group was also highly enriched for cardiovascular endpoints such as heart failure (I50.9, ICD-10), atrial fibrillation and flutter (I48, ICD-10), and acute myocardial infarction (I21*, ICD-10).

### PheWAS

We observed several phenome-wide associations across adverse events, BP traits, and clinical endpoints (**Supplementary Tables 7 –10**). In the adverse event PheWAS, 13/21 ACEI Switch CS lead variants showed at least nominal associations with cough after ACEI initiation (**Figure 5**). We did not observe statistically significant associations between dCCB switching variants and local oedema (**Supplementary Table 8**).

**Figure 5.**
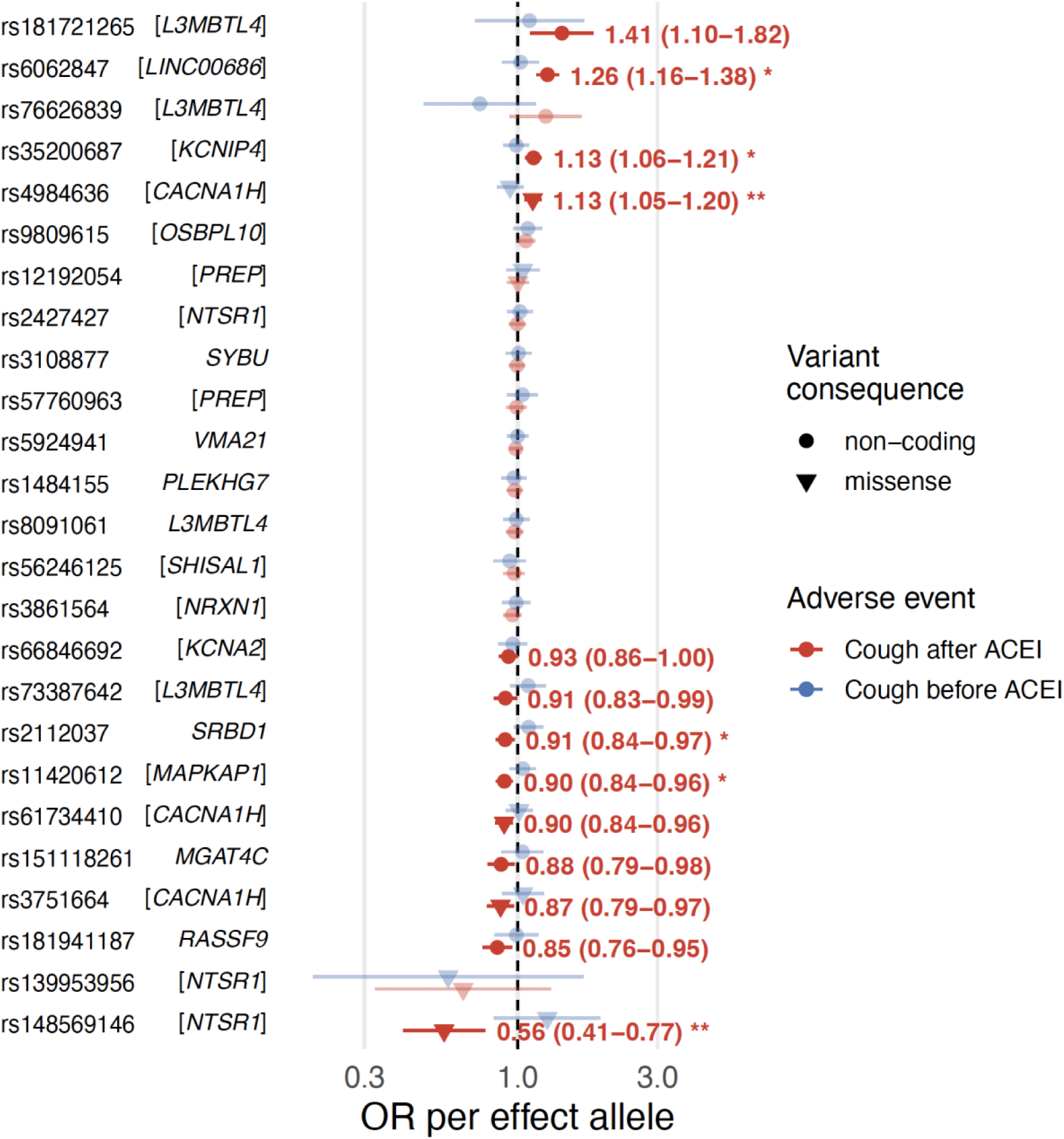
Associations between ACEI Switch variants and cough risk before and after ACEI initiation. Odds ratios (OR) and 95% confidence intervals per effect allele are shown among all ACEI users in FinnGen (N=93,283), separately for one-year periods before (blue) and after (red) ACEI initiation. Gene-drug interactions were tested with conditional fixed-effects logistic regression in a self-controlled design. Asterisks denote nominally (*P<0.05) or Bonferroni-significant (**P<0.05/25) interactions. Missense and non-coding variants are shown as filled triangles and circles, respectively. Shapes are faded when the 95% confidence interval overlaps with the dashed line (OR=1). Brackets around gene names indicate that the variant is inside the gene. ACEI, angiotensin-converting enzyme inhibitor.

In the BP GWAS by Keaton et al., only the Switch-protective CS lead variant rs11420612-AG in *MAPKAP1* was associated with BP traits: systolic BP (beta=-0.18, P=2.0×10^−11^), pulse pressure (beta=-0.10, P=9.3×10^−8^), and diastolic BP (beta=-0.08, P=6.0×10^−7^) (**Supplementary Table 9**). In the clinical endpoint PheWAS, we observed positive associations with hypertension (*CACNA1H* p.Val664Ala), antihypertensive use (*CACNA1H* p.Val664Ala, *CYP3A43*), and cough (*LINC00686*, *KCNIP4*); several negative sleep and respiratory tract-related associations for *MAPKAP1*, and a positive episodal and paroxysmal disorder association for *CACNA1H* p.Val664Ala (**Supplementary Table 10**).

### Drug-gene interaction analysis

In a self-controlled risk interval analysis for ACEI exposure using conditional logistic regression, we observed statistically significant drug-gene interactions for missense variants *NTSR1* p.Gly301Arg (interaction OR=0.39, P=8.1×10^−4^) and *CACNA1H* p.Val664Ala (interaction OR=1.25, P=1.2×10^−3^) after Bonferroni correction (P<0.05/25), and nominal (P<0.05) interactions for CS lead variants in *LINC00686*, *KCNIP4*, *MAPKAP1*, and near *SRBD1* (**Figure 5**, **Supplementary Table 11**).

## Discussion

We mapped the genetic architecture of short-term antihypertensive treatment failure in three large population-based biobanks (FinnGen, the Estonian Biobank and the UK Biobank) including over 400,000 individuals prescribed antihypertensives. By leveraging switching patterns across major antihypertensive classes, we identified novel pharmacogenetic loci that implicate medication class-specific pathways underlying treatment failure.

Multiple independent analyses strongly implicate the neurotensin pathway in ACEI-induced cough pathophysiology. First, we identified two independent neurotensin receptor gene *NTSR1* missense variants, p.Gly301Arg and p.Tyr339Cys, with concordant effects on switching from ACEI (**Figure 2**). Second, the strongest ACEI Switch signal at the 40-fold Finnish-enriched rs181941187-G allele near *RASSF9* tags a cluster of variants near the neurotensin gene *NTS* (**Figure 3**). Third, we demonstrated a gene-drug interaction whereby the *NTSR1* p.Gly301Arg decreases ACEI-associated cough risk to pre-exposure levels (**Figure 5**). Together, these findings motivate a reinterpretation of ACEI cough biology in the context of the neurotensin pathway.

Accumulation of bradykinin and substance P during ACE inhibition remains the widely accepted explanation for ACEI-induced cough.[28] However, candidate gene studies of bradykinin pathway genes such as *BDKRB2* and *ACE* have yielded inconsistent results,[29] and neither bradykinin nor substance P has been implicated in genome-wide studies of ACEI intolerance [6,7,30]. Interestingly, neurotensin and substance P share similar biological functions,[31] including degradation by ACE.[32] Therefore, neurotensin accumulation is also expected during ACE inhibition, although *in vivo* evidence of this phenomenon is limited. We propose to expand the classical bradykinin-substance P framework to include neurotensin signaling, arguing that it is genetically and mechanistically supported.

Duan et al. show that the NTSR1 glycine residue 301^6.33^ (Ballesteros-Weinstein numbering) forms part of a docking interface for signal transducers,[33] and Zhou et al. predict it to participate in G protein coupling.[34] Substitution of the small, uncharged glycine with a much larger, positively charged arginine (i.e. p.Gly301Arg) could plausibly shift downstream signaling by modifying receptor-transducer interactions. Although no direct functional data exist for this variant, experimental data in homologous human receptors provide mechanistic evidence. In recent papers, Liao and Yue demonstrate that mutations at corresponding 6.33 positions – L341^6.33^A in the cannabinoid receptor 1 and L246^6.33^A in the apelin receptor – induce more than tenfold biased signaling toward beta-arrestin 1.[35,36] Recent rodent studies link such beta-arrestin biased NTSR1 signaling to reduced addiction-related behaviour and reduced pain sensitivity.[37,38] Intriguingly, p.Gly301Arg was nominally associated with lower all-cause pain in FinnGen (OR=0.93, P=6.6×10^−5^), consistent with these experimental findings (**Supplementary Table 10**). Biased modulation is currently being pursued in next-generation NTSR1 drug development.[39] Based on this mechanistic evidence, the overlap with known ACEI biology, and the established link between chronic dry cough and pain signaling,[7] we hypothesize that p.Gly301Arg acts as a constitutive biased modulator of NTSR1 and exerts prominent cough-protective effects under ACEI inhibition.

Our work extends previous research on non-coding genetic variation underlying ACEI intolerance. Ghouse et al. performed an ACEI-to-ARB switching GWAS meta-analysis across 3 cohorts (33,959 cases, 44,041 controls) and identified 7 genes: *KCNA2*, *SRBD1*, *KCNIP4*, *PREP*, *SCAI*, *L3MBTL4*, and *NTSR1*.[6] We re-discover and replicate all but *SCAI*. Coley et al. studied the genetic overlap between ACEI-to-ARB switching (21,523 cases, 62,569 controls) and chronic dry cough (7635 cases, 88,653 controls), identifying 14 loci mapping to 19 genes: *VMA21*, *CYP27B1*, *PREP*, *NTSR1*, *KCNA10*, *OR4C12*, *OR4C13*, *KCNIP4*, *MAPKAP1*, *CTNNA1*, *SIL1*, *RASSF9*, *SLCO4A1*, *L3MBTL4*, *SRBD1*, *ALX1*, *RBM15*, *CPEB2*, and *ATP23*.[7] We replicate 9 of these genes. Our novel ACEI switching loci at *SYBU*, *CACNA1H*, and *SHISAL1* broaden the polygenic spectrum of ACEI intolerance. *SYBU* encodes a kinesin adaptor for axonal cargo and mitochondria, *CACNA1H* encodes the Cav3.2 T-type calcium-channel α1H subunit, and *SHISAL1* is a membrane protein mainly expressed in neuronal tissue. Together, these findings reinforce the emerging view that neural excitability pathways underlie ACEI intolerance.

The CYP3A4 enzyme metabolizes approximately half of all clinically prescribed medications, including dCCB.[40] Our externally replicated dCCB Switch lead variant is in near-complete linkage with *CYP3A4*22*, a recently characterized pharmacogenetic variant associated with reduced CYP3A4 enzyme activity[40] and higher dCCB serum concentrations.[41] Elevated dCCB levels increase the risk of side-effects such as peripheral oedema, which may lead to medication switching. We report a robust association between *CYP3A4*22* and real-world dCCB switching across FinnGen, UKB, and EstBB. These novel findings provide genome-wide, cross-biobank evidence supporting genotype-informed antihypertensive medication selection in *CYP3A4*22* carriers.

We observed contrasting genetic architectures between Switch and Discontinue phenotypes across major antihypertensive classes (**Figure 4**). Discontinue phenotypes were highly correlated, implying a shared genetic basis of antihypertensive discontinuation across medication classes, which aligns with Cordioli et al. who identified strong hypertension loci as drivers of antihypertensive use persistence.[9] However, such a pattern was absent among Switch phenotypes. Moreover, the consistent attenuation of genetic effect sizes between ACEI switching and discontinuation suggests strong confounding in the Discontinuation phenotype, possibly driven by poor persistence unrelated to medication response (**Supplementary Figure 4**). Although the heritabilities of antihypertensive use patterns were relatively low, this does not preclude pharmacogenetic discovery, as demonstrated in this study. We conclude that subtyping treatment failure into switching and discontinuing can increase phenotype specificity and improve statistical power in biobank-scale pharmacogenetic studies, as shown here.

The results of this study should be viewed in the context of its limitations. First, neither medication purchases in FinnGen and EstBB nor prescriptions in UKB guarantee medication use. If a large proportion of individuals did not use their medications, the switching phenotypes may reflect patient characteristics other than drug response. However, repeated purchases such as in Switch and Continue phenotypes likely indicate true underlying medication use. Second, we were unable to directly observe the underlying reasons for switching or discontinuation. Finally, the focus on Finnish and European ancestry limits the generalizability of our findings to other ancestries.

In conclusion, our findings implicate *NTSR1* p.Gly301Arg and the neurotensin signaling pathway as pharmacogenetically and mechanistically supported contributors to ACEI-induced cough, identify *CYP3A4*22* as a risk-increasing variant for dCCB intolerance, and underscore the value of medication use patterns for pharmacogenetic discovery. Together, these results demonstrate how biobank-scale longitudinal medication data can help advance the emerging field of statistical pharmacogenomics.

## Acknowledgements

We want to acknowledge the participants and investigators of the FinnGen study. The FinnGen project is funded by two grants from Business Finland (HUS 4685/31/2016 and UH 4386/31/2016) and the following industry partners: AbbVie Inc., Alnylam Pharmaceuticals, Inc., AstraZeneca UK Ltd, Bayer AG, Biogen MA Inc., Boehringer Ingelheim International GmbH, Bristol Myers Squibb Inc. (and Celgene Corporation & Celgene International II Sàrl), Genentech Inc., GlaxoSmithKline Intellectual Property Development Ltd., Johnson&Johnson Innovative Medicine Inc., Maze Therapeutics Inc., Merck Sharp & Dohme LCC, Novartis AG, Pfizer Inc. and Sanofi US Services Inc. Following biobanks are acknowledged for delivering biobank samples to FinnGen: Auria Biobank (www.auria.fi/biopankki), THL Biobank (www.thl.fi/biobank), Helsinki Biobank (www.helsinginbiopankki.fi), Biobank Borealis of Northern Finland (https://www.ppshp.fi/Tutkimus-ja-opetus/Biopankki/Pages/Biobank-Borealis-briefly-in-English.aspx), Finnish Clinical Biobank Tampere (www.tays.fi/en-US/Research_and_development/Finnish_Clinical_Biobank_Tampere), Biobank of Eastern Finland (www.ita-suomenbiopankki.fi/en), Central Finland Biobank (www.ksshp.fi/fi-FI/Potilaalle/Biopankki), Finnish Red Cross Blood Service Biobank (www.veripalvelu.fi/verenluovutus/biopankkitoiminta), Terveystalo Biobank (www.terveystalo.com/fi/Yritystietoa/Terveystalo-Biopankki/Biopankki/) and Arctic Biobank (https://www.oulu.fi/en/university/faculties-and-units/faculty-medicine/northern-finland-birth-cohorts-and-arctic-biobank). All Finnish Biobanks are members of BBMRI.fi infrastructure (https://www.bbmri-eric.eu/national-nodes/finland/). Finnish Biobank Cooperative -FINBB (https://finbb.fi/) is the coordinator of BBMRI-ERIC operations in Finland. The Finnish biobank data can be accessed through the Fingenious® services (https://site.fingenious.fi/en/) managed by FINBB. We acknowledge all the participants of the Estonian Biobank. Data analysis was carried out in part in the High-Performance Computing Center of the University of Tartu. The research was conducted using the Estonian Center of Genomics/Roadmap II funded by the Estonian Research Council (project number TT17). We acknowledge all participants of the UK Biobank. This research has been conducted using the UK Biobank Resource under Application Number 22627.

## Data availability statement

FinnGen data access is described at https://www.finbb.fi. UK Biobank data access is described at https://www.ukbiobank.ac.uk/enable-your-research. Estonian Biobank data access is described at https://genomics.ut.ee/en/content/estonian-biobank#dataaccess.

