## Supplementary Figures 1-6 for "Pharmacogenomic architecture of antihypertensive switching implicates neurotensin-NTSR1 signaling in ACE inhibitor-induced cough"

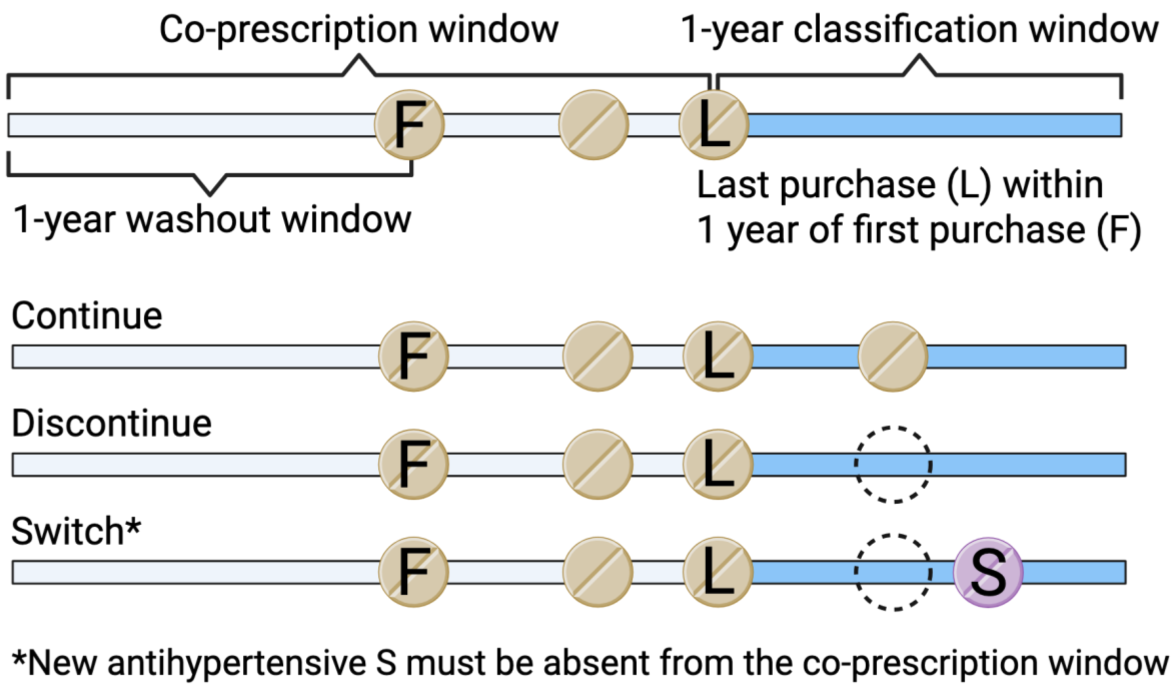


**Supplementary Figure 1. Medication use pattern phenotyping.** For a given index medication class (e.g. angiotensin-converting enzyme inhibitors) and index substance within the index class (e.g. enalapril), each individual's medication history was divided into three partially overlapping windows: a 1-year washout window preceding the first index class purchase (F) to ensure sufficient follow-up, a co-prescription window extending from the beginning of the washout window to the last purchase (L) within one year of F, and a subsequent 1-year classification window starting from L. Participants were classified as Continue if the classification window contained at least one purchase of the same index substance (e.g. enalapril), Discontinue if no purchases of the index substance (e.g. enalapril) occurred, and Switch if a new antihypertensive substance (S) absent from the co-prescription window was purchased in the classification window. We allowed S to be from the index class (e.g. ramipril) to allow for both inter-class and intra-class switching. We also allowed S to be from outside the five major antihypertensive classes. Created with BioRender.com.

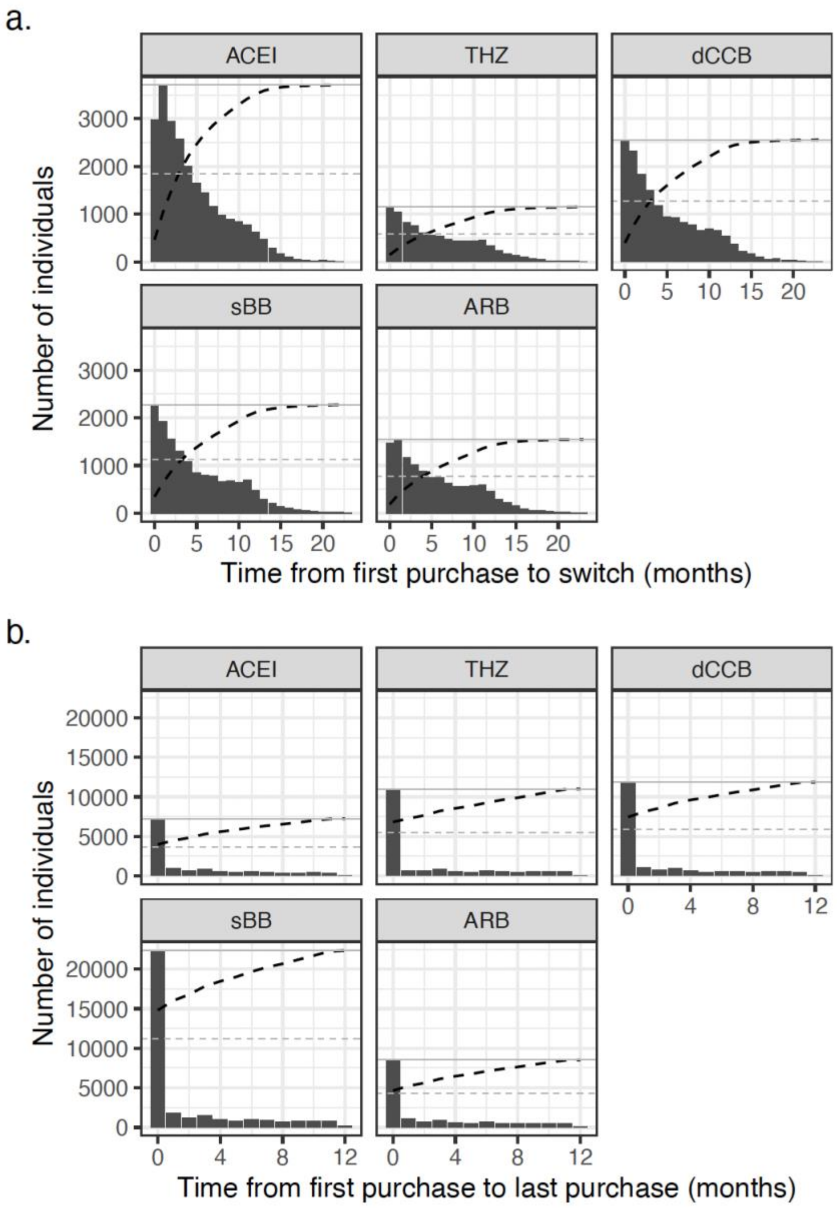


**Supplementary Figure 2. Time to switching and discontinuation across antihypertensive classes.** (a) Distribution of time in months from first purchase to new antihypertensive medication among participants classified as Switch. (b) Distribution of time in months from first purchase to last purchase (can be the same as first purchase) among participants classified as Discontinue. ACEI, angiotensin-converting enzyme inhibitor; ARB, angiotensin 2 receptor blocker; dCCB, dihydropyridine calcium channel blocker; sBB, selective beta blocker; THZ, thiazide diuretic. Dashed black line indicates cumulative proportion, dashed gray line 50%, and solid gray line 100%.

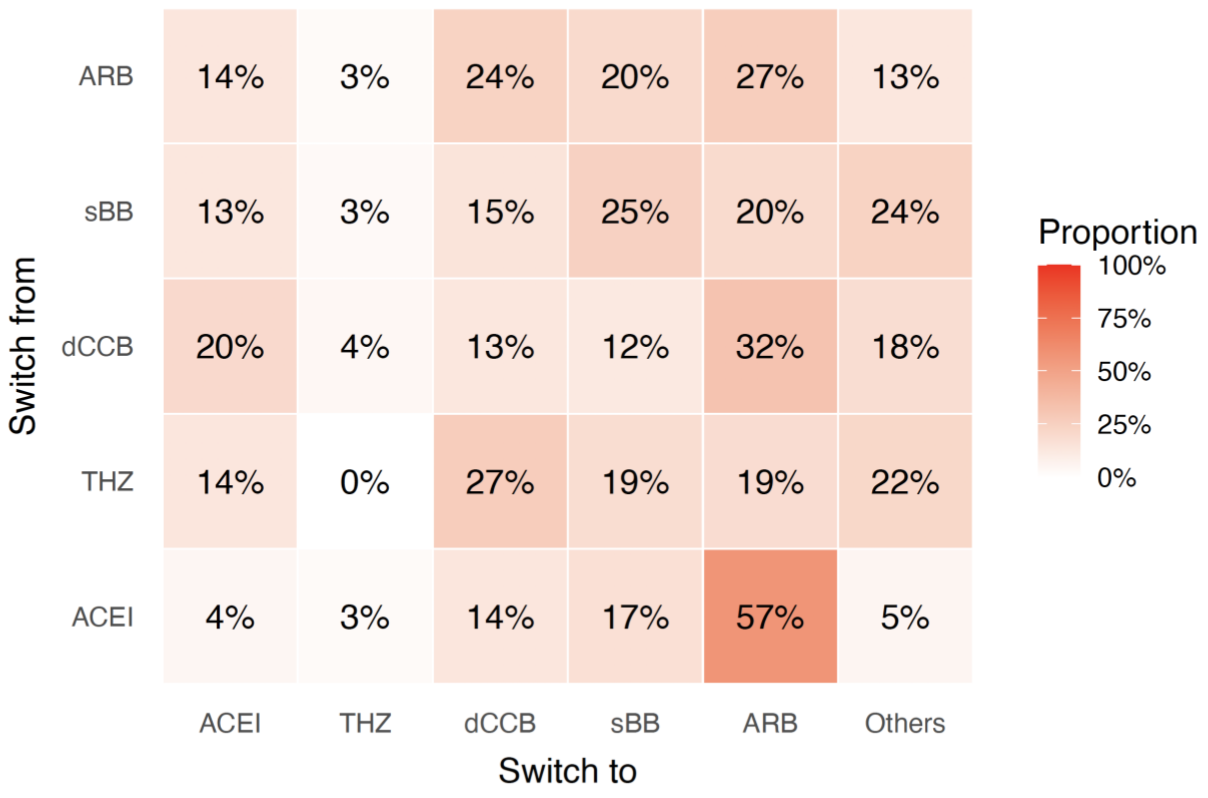


**Supplementary Figure 3. Heatmap of switching patterns among antihypertensive users.** Heatmap showing the proportion of individuals switching from each antihypertensive class (rows) to another medication in the same or different antihypertensive class (columns). The heatmap includes only individuals classified as Switch. ACEI, angiotensin-converting enzyme inhibitor; ARB, angiotensin 2 receptor blocker; dCCB, dihydropyridine calcium channel blocker; sBB, selective beta blocker; THZ, thiazide diuretic.

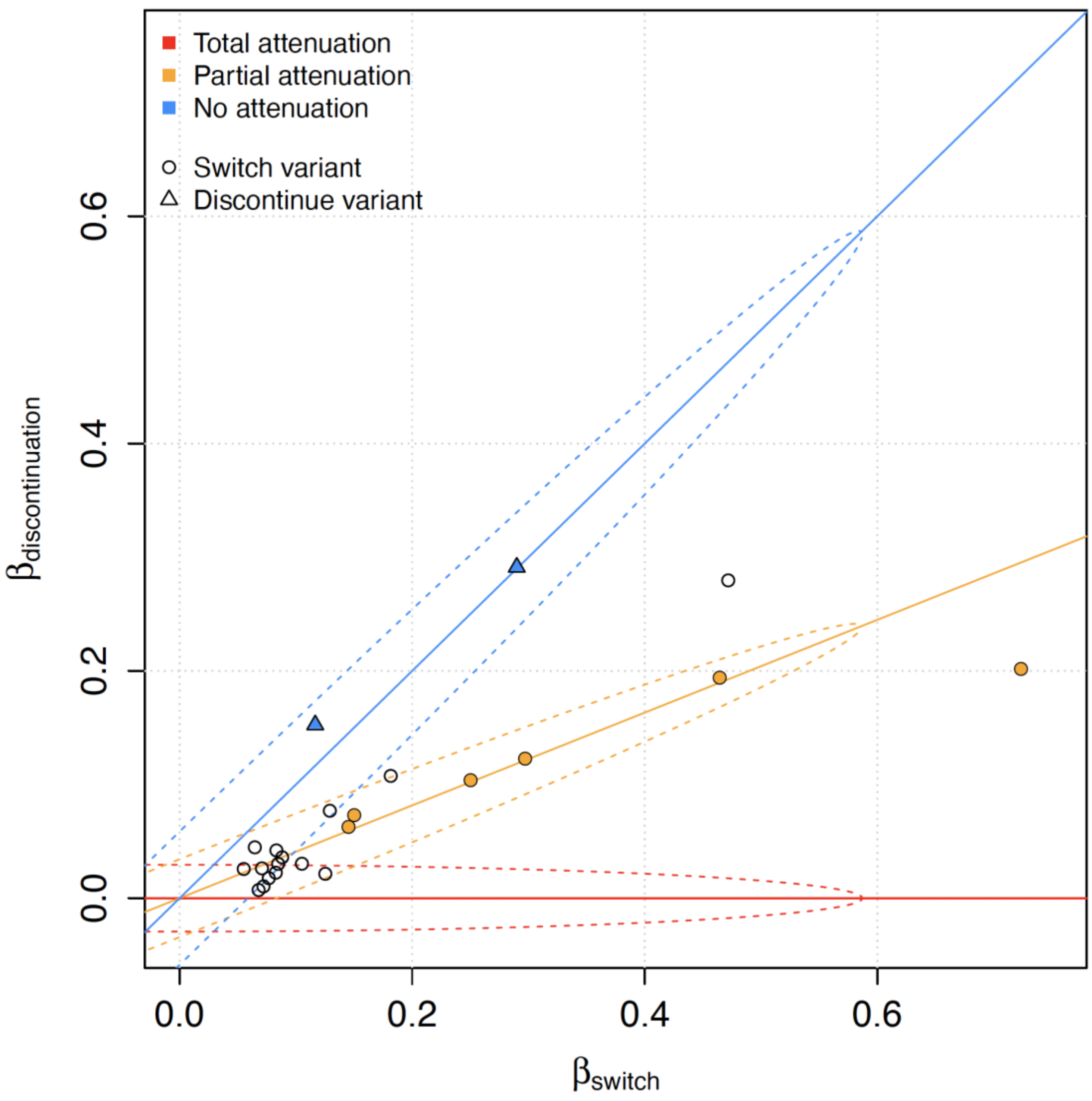


**Supplementary Figure 4. Asymmetric attenuation of genetic effects between ACEI Switch and Discontinue phenotypes in FinnGen.** Each point shows the effect size of a lead variant in ACEI Switch (x-axis) and Discontinue (y-axis), with circles denoting Switch variants and triangles Discontinue variants. Reference lines indicate attenuation models corresponding to total attenuation (red, slope=0), partial attenuation (yellow, slope=0.41), and no attenuation (blue, slope=1) in the Discontinue phenotype. Switch variants cluster along the partial attenuation model, indicating marked attenuation of their effects in the Discontinue phenotype. In contrast, Discontinue variants cluster along the no attenuation model, indicating unattenuated transfer of effects from Discontinue to Switch. Coloured shapes indicate >95% posterior probability of model classification. ACEI, angiotensin-converting enzyme inhibitor.

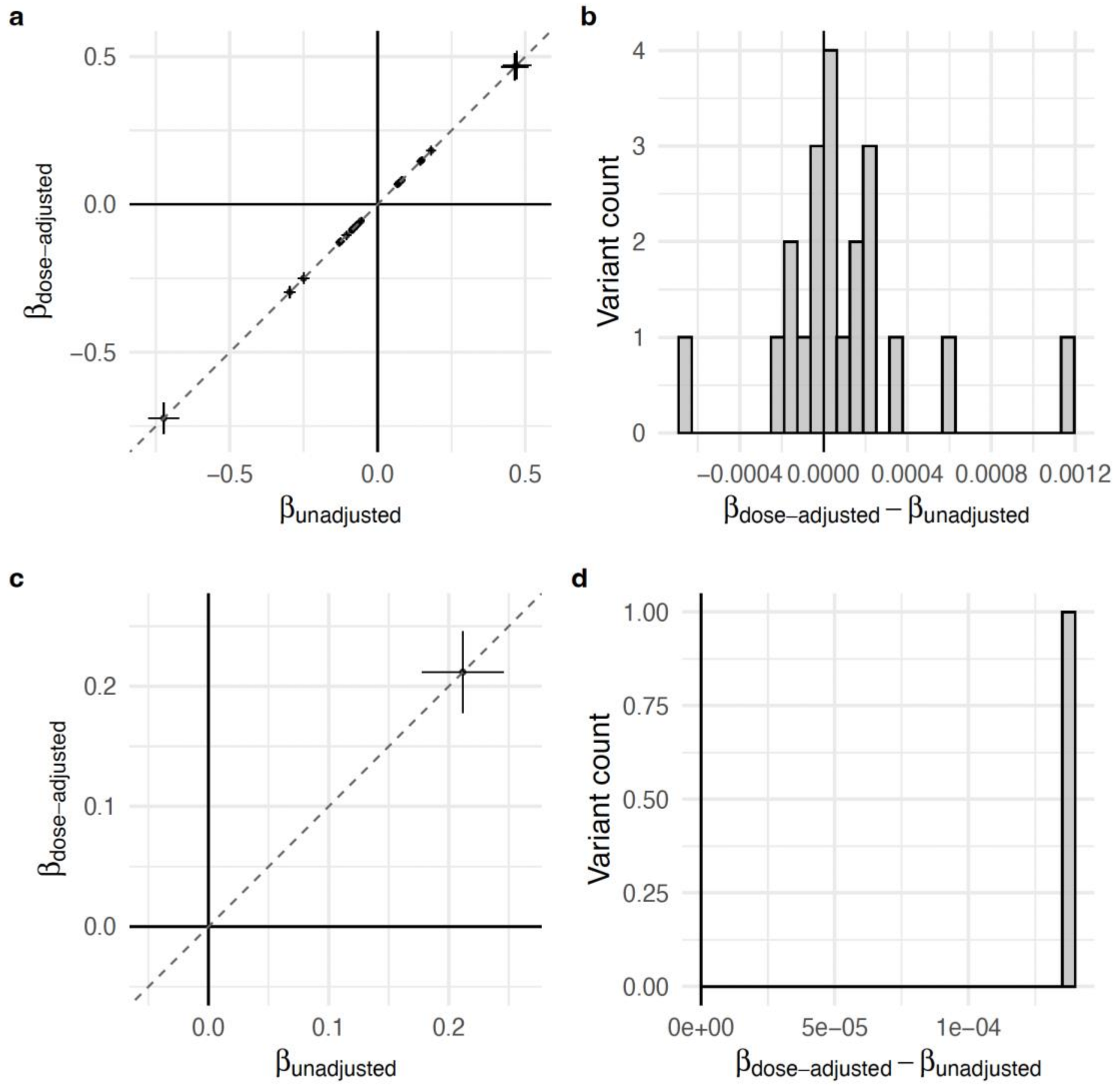


**Supplementary Figure 5. Comparison of unadjusted and dose-adjusted effect size estimates for ACEI (a, b) and dCCB (c, d) Switch credible set lead variants.** Panels (a, c) show dose-adjusted versus unadjusted β estimates, while panels (b, d) show the distribution of their arithmetic differences. Effect sizes were consistent across models.

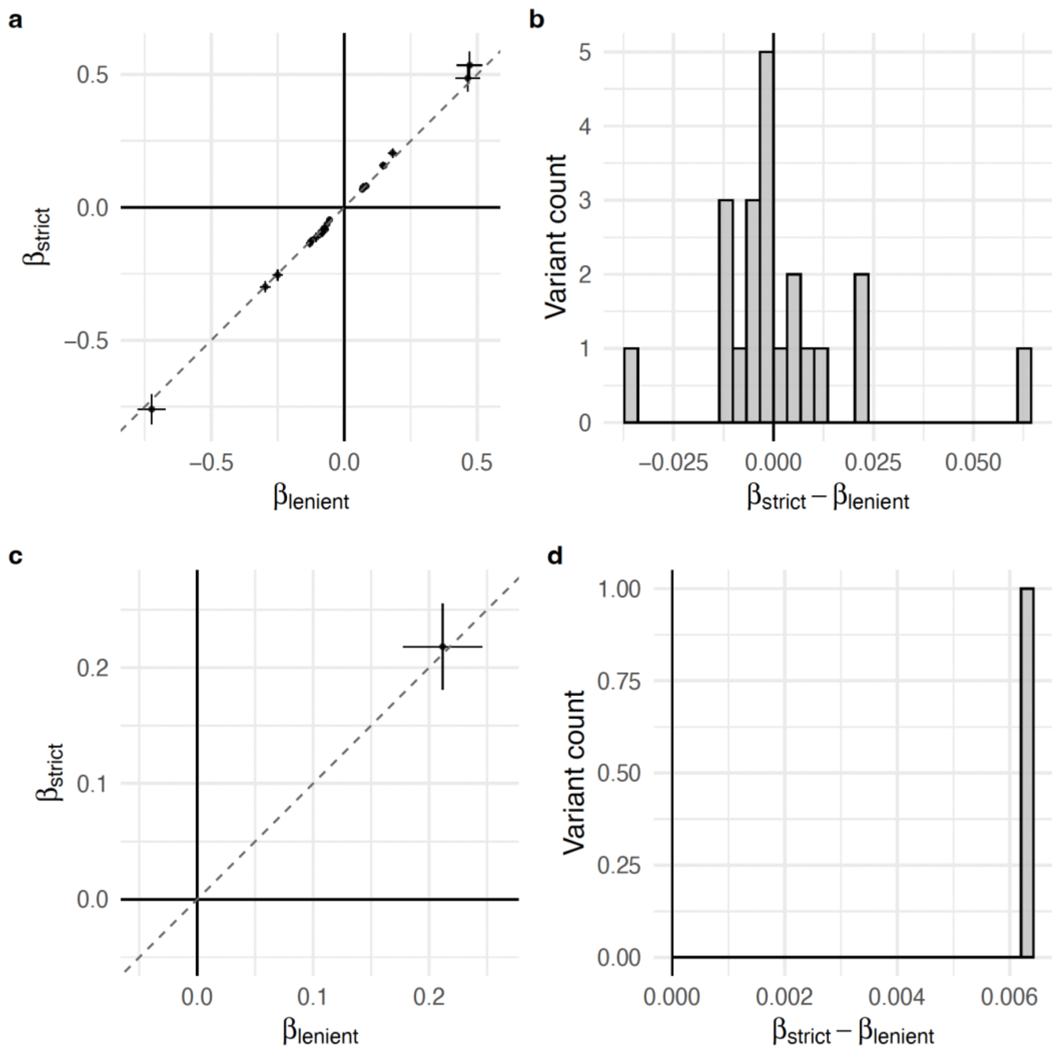


**Supplementary Figure 6. Comparison of unadjusted and strict-adherence effect size estimates for ACEI (a, b) and dCCB (c, d) Switch credible set lead variants.** Panels (a, c) show effect sizes for strict adherence (y-axis, max 180 days between purchases) versus lenient adherence (x-axis), while panels (b, d) show the distribution of their arithmetic differences. Effect size estimates were consistent across adherence definitions.
